# Human-centred design approaches to health facility design: Evidence from perinatal care settings in Ethiopia and Bangladesh

**DOI:** 10.64898/2026.06.05.26354949

**Authors:** Stephen Luna-Muse, Minara Chowdhury, Raihan Sharif, Sonya Panjwani Olaya, Johanna Figueroa, Amie Shao, Andrew Brose, Mikail Jassat, Pierre Barker

## Abstract

While significant progress has been made in perinatal outcomes over recent decades in low- and middle-income countries (LMICs), maternal and newborn quality improvement initiatives often fail to account for the spatial conditions in which they are implemented. Health systems are increasingly deploying evidence-based care models into built environments that are not optimally structured to meet the needs of its patient population. As the principal users, patients and health care workers can offer pragmatic insights about improving these structural designs. Our objective was to gather insights from patients, providers, and companions about how the physical design of their health facilities influenced their experience receiving or delivering perinatal care. We conducted a prospective observational study using a human-centred design (HCD) approach to analyse perceptions of the quality of perinatal care across two low resource settings: Ethiopia and Bangladesh. Using engagement and assessment tools, we conducted interviews, focus groups, facility walk-throughs, co-design workshops, and infrastructural assessments with patients, companions, providers, and Ministry of Health representatives. Descriptive statistics and thematic analysis were used to identify key learnings and develop recommendations. Across both countries, participants identified the need for facility layouts that better support privacy, mobility during labour, alternative birth positions, companion involvement, cultural and religious practices, sanitation, and provider visibility. Based on these insights, we developed six recommendations to better align health facility infrastructure with maternal and newborn care delivery needs. Our findings suggest that investments in health facility infrastructure may improve care experiences and help enable respectful, safe, and evidence-based maternal and newborn care. Alongside targeted spatial improvements, government authorities responsible for health facility planning should incorporate participatory design processes to ensure infrastructure reflects the needs of patients, companions, and providers and supports high-quality care delivery.

## Introduction

In recent decades, substantial progress in perinatal health has been driven by health systems strengthening efforts, expanded antenatal care, epidemiological advances, and reductions in HIV/AIDS mortality (Cresswell et al., 2025). However, far less attention has been paid to whether the physical environments in which care is delivered adequately support recommended models of maternal and newborn care. Recent studies suggest that poorly designed care environments across the perinatal care continuum are strongly associated with poorer health outcomes and experiences (Ban et al., 2021; Goldkuhl et al., 2023; Setola et al., 2019).

Several studies in low- and middle-income countries (LMICs), including Ethiopia and Zambia, identify clinical environments as potential barriers to – or enablers of – facility-based childbirth (Scott et al., 2018; Sialubanje et al., 2015; Stierman et al., 2023). In Zambia, lack of space in the maternity wards, poor clinical infrastructure (no water, electricity, and cell network), and no beds were cited as top problems shared by pregnant women during their delivery (Scott et al., 2018). Another systematic review of barriers to obstetric care in sub-Saharan Africa similarly highlighted inadequate infrastructure and overcrowding as barriers to receiving quality care (Kyei-Nimakoh et al., 2017). These studies collectively underscore the critical need to align health facility infrastructure with the operational requirements of respectful, safe, and evidence-based perinatal care.

Improving health facility infrastructure also aligns with the WHO Standards for Improving Quality of Maternal and Newborn Care in Health Facilities, particularly Standard 5: Women and newborns receive care with respect and preservation of their dignity and Standard 8: The health facility has an appropriate physical environment, with adequate water, sanitation and energy supplies, medicines, supplies and equipment for routine maternal and newborn care and management of complications (World Health Organization, 2016). Despite growing emphasis on respectful and evidence-based maternal and newborn care, the spatial and infrastructural requirements needed to implement these models are rarely incorporated into facility planning processes.

### Context and Study Settings

The study was undertaken in clinical facilities in Ethiopia and Bangladesh.

In Ethiopia, only half of mothers give birth under the supervision of a Skilled Birth Attendant, the majority of those taking place in health facilities (Mekuria et al., 2025); a similar proportion (53%) of births in Bangladesh are attended by a skilled provider (Afroja et al., 2022). In studies conducted to date, women have stated their aversion to delivering in facilities is due to lack of privacy (e.g., overlap of labouring and postpartum mothers in the same room) and poor layout of labour and delivery rooms, making it difficult for providers to safely care for high volumes of patients and for patients to receive privacy and companionship, among other challenges (Kyei-Nimakoh et al., 2017).

Bangladesh has seen a notable reduction (79%) in maternal mortality; however, disparities persist (UNICEF, 2025). Nearly half of births occur at home, with a higher percentage of home births in rural areas (Shibre et al., 2021). Recent studies have indicated women in Bangladesh choose not to seek perinatal care due to structural barriers, including cleanliness concerns and lack of privacy, among other concerns (Chanda et al., 2025; “Unseen Obstacles,” 2025). General hospitals at district level are currently designed, constructed, and maintained by engineers from the Ministry of Health and Family Welfare Infrastructure Directorate’s Public Works Department, while the Health Engineering Department leads design, construction, and maintenance of Upazila Health Complexes (subdistrict level hospitals). While these agencies follow strict standards to ensure buildings are safe, maternal and newborn services are often delivered in spaces designed without consideration of local care practices, patient flow, companionship, privacy, or provider workflow needs.

Our study sought to understand how the physical structure and spatial organization of health facilities impacts the delivery and experience of perinatal care in Bangladesh and Ethiopia, and how gaps between care models and facility infrastructure may constrain implementation of respectful, patient-centred maternal and newborn care. Using a human-centred design (HCD) approach, we sought to propose spatial and infrastructural improvements that better support care delivery and improve experiences for mothers, companions, and providers.

## Methods

### Study Design

Between August and November 2021, we conducted a prospective observational study using a HCD approach to examine perceptions and experiences of perinatal care across two geographically and culturally distinct settings, Ethiopia and Bangladesh. A HCD approach was selected because it incorporates ethnographic and observational methods to understand the values and needs of stakeholders and develop contextually aligned improvements through a structured and iterative process (Roberts et al., 2016; Zuber & Moody, 2018).

### Study Sample

In partnership with the Ministries of Health in each country, we used purposive sampling to select four facilities per country, two hospitals and two health centres. Facilities were selected based on their representativeness of similar facilities within the health system, opportunities for replication across settings, available space, and potential for future expansion. A description of each facility is included in Table 1.

**Table 1.** Study setting characteristics.

|  | **Hospital A**<br>**(Munshiganj**<br>**General**<br>**Hospital)** | **Hospital B**<br>**(Jashore**<br>**General**<br>**Hospital)** | **Hospital C**<br>**(Addis**<br>**Zemen**<br>**Primary**<br>**Hospital)** | **Hospital D**<br>**(Mohammad**<br>**Akle**<br>**General**<br>**Hospital)** | **Health**<br>**Centre A**<br>**(Serajdikhan**<br>**Upazila**<br>**Health**<br>**Complex)** | **Health**<br>**Centre B**<br>**(Abhaynagar**<br>**Upazila**<br>**Health**<br>**Complex)** | **Health**<br>**Centre C**<br>**(Yifag**<br>**Health**<br>**Centre)** | **Health**<br>**Centre D**<br>**(Melka**<br>**Worer**<br>**Health**<br>**Centre)** |
| --- | --- | --- | --- | --- | --- | --- | --- | --- |
| **Country** | Bangladesh | Bangladesh | Ethiopia | Ethiopia | Bangladesh | Bangladesh | Ethiopia | Ethiopia |
| **District** | Dhaka | Khulna | Amhara | Afar | Dhaka | Khulna | Amhara | Afar |
| **Urban city** | Urban | Urban | Rural | Rural | Rural | Rural | Rural | Rural |
| **Scope of**<br>**Services** | Antenatal,<br>labour and<br>delivery,<br>obstetrics<br>and<br>gynecology,<br>postnatal | Antenatal,<br>labour and<br>delivery,<br>postnatal,<br>neonatal<br>intensive<br>care unit | Family<br>planning,<br>antenatal,<br>labour and<br>delivery,<br>postnatal,<br>neonatal<br>intensive<br>care unit,<br>outpatient<br>department,<br>operating<br>room | Family<br>planning,<br>including<br>abortion<br>care,<br>antenatal,<br>labour and<br>delivery,<br>postnatal,<br>neonatal<br>intensive<br>care unit,<br>outpatient<br>department,<br>operating<br>room | Family<br>planning<br>including<br>antenatal,<br>labour and<br>delivery,<br>postnatal<br>outpatient<br>department | Antenatal,<br>labour and<br>delivery,<br>postnatal | Antenatal,<br>labour and<br>delivery,<br>postnatal,<br>outpatient<br>department | Family<br>planning,<br>antenatal,<br>labour<br>and<br>delivery,<br>postnatal |
| **Patient**<br>**Volume** | 1269 births<br>per year | 1847 births<br>per year | 1972 births<br>per year | 752 births<br>per year | 124 births<br>per year | 536 births per<br>year | 266 births<br>per year |  |

### Data Collection

Data collection tools were co-designed with local partners in Ethiopia and Bangladesh to capture experiences of childbirth care and assess the physical structure and condition of health facilities and surrounding environments. Both tools were reviewed by a gender advisor to ensure they reflected considerations of dignity, autonomy, and privacy.

Participants included mothers who had delivered within the preceding three months, family members or companions, health care providers who had worked at the facility for at least six months, community members, and Ministry of Health representatives. Purposive sampling was used for interviews and focus groups, while convenience sampling was used for quantitative surveys with postpartum mothers.

Data collection included semi-structured interviews, focus groups, co-design workshops, facility walk-throughs, observational shadowing, and care-flow simulations with mothers, companions, providers, administrators, and community members. Field notes, reflection sheets, photographs, and audio/video recordings (with participant consent) were used to document observations and support analysis.

A second team of architects and engineers conducted infrastructural assessments of each facility, including evaluations of building condition, water and sanitation systems, electrical systems, and spatial organization. Working alongside providers, the team mapped patient care flows and documented how patients moved through perinatal care spaces. A summary of engagement activities conducted in each setting is shown in Table 2.

**Table 2.** Data collection in each setting.

| Activity | # Completed |  |  |  |
| --- | --- | --- | --- | --- |
|  | Amibara District, Ethiopia | Libokemkem District, Ethiopia | Munshiganj District, Bangladesh | Jashore District, Bangladesh |
| **Mothers in Facility** |  |  |  |  |
| Semi-structured interview with postpartum mothers | 6 | 4 | 12 | 8 |
| Semi-structured interview with mothers of small and sick newborns (SSNB) | 2 | 2 | 0 | 4 |
| Quantitative survey for mothers | 21 | 22 | 8 | 8 |
| Facility walk-through with mothers | 1 | 2 | 2 | 2 |
| Observation/shadowing of mothers | 2 | 2 | 2 | 2 |
| **Family / Companions in Facility** |  |  |  |  |
| Semi-structured interview with family / companions | 5 | 6 | 4 | 4 |
| **Providers in Facility** |  |  |  |  |
| Introductory interview and facility tour with administrator and provider | 2 | 2 | 2 | 2 |
| Semi-structured interview with providers | 7 | 4 | 8 | 8 |
| Experience simulation with providers (*in which an Immersion Team member acted as a pregnant mother and clinicians simulated the care they would provide*) | 2 | 2 | 2 | 2 |
| Co-design workshop for providers | 2 | 2 | 2 | 2 |
| Space use documentation with administrator or provider | 2 | 2 | 2 | 2 |
| **Community** |  |  |  |  |
| Semi-structured interview for recently delivered mothers | 2 | 1 | 8 | 8 |
| Focus group workshop for recently delivered mothers | 1 | 2 | 2 | 2 |
| Birth experience simulation in community | 0 | 0 | 2 | 2 |
| **Ministry of Health (MOH / RHB)** |  |  |  |  |
| Interview guide for Maternal-Newborn Health | 1 | 1 | 1 | 1 |
| Interview guide for Infrastructure Directorate | 2 | 1 | 1 | 1 |

### Data Analysis

Following data collection, interviews and observation notes were transcribed, organized, and analysed. Qualitative data were organized by respondent type and data collection method to support comparative thematic analysis across stakeholder groups and settings. Qualitative data were analysed inductively using thematic analysis.

### Data Synthesis

The project teams’ architects and engineers used the data analysis to inform draft concept designs proposing spatial improvements to the participating facilities to better meet the needs of both patients and providers. These concepts were iteratively reviewed with facility leadership, Ministries of Health, and other relevant stakeholders.

In addition to site-specific concept designs, the team developed a set of cross-cutting design principles addressing recurring concerns across both countries, including care flow adjacency, overcrowding, privacy, companion accommodation, and mobility during labour.

### Ethics Review

In Ethiopia, the study protocol was approved by the Ethiopian Public Health Association Institutional Review Board [IRB Number: EPHA-OG-XXX-XX; Approval Date: 28 April 2021]. The study protocol was approved by the International Centre for Diarrhoeal Disease Research Ethics Review Committee in Bangladesh [Research Protocol Number: PR-XXXXX; Approval Date: 1 August 2021].

## Results

Thematic analysis of the interviews, surveys, focus groups, and facility walk-throughs across all eight facilities in Bangladesh and Ethiopia resulted in six themes related to overall facility and space layout, companion accommodation, cultural and religious practices, water and sanitation facilities, mobility during labour and alternative birth positions, and provider proximity and visibility (see Table 3 for summary of key themes).

**Table 3.** Summary of Key Themes.

| Category | Key Themes | Sample Quote |
| --- | --- | --- |
| **Overall Space & Adjacencies** | - Space constraints due to budget, poor planning, and lack of engagement with users<br>- Crowded facilities leading to congestion, discomfort, and privacy concerns<br>- Inefficient layouts disrupt patient flow and privacy | *"[There are] two or three patients and their attendants are present in one room. All examination and delivery is done in the same room. So, there is no privacy."* |
| **Companions** | - Essential role in supporting mothers, yet their needs are unable to be accommodated<br>- Lack of seating, sleeping, and storage facilities forces them into overcrowded areas | *"I have been sleeping outside the MNH block on a piece of cardboard because there is no other option while we stay here."* |
| **Cultural Practices** | - Religious and cultural practices are an important component during the birth experience<br>- Facilities have difficulties accommodating traditional food preparation, religious rites, and prayer spaces<br>- Lack of potable water further limits hygiene and cultural observances | *"[We'd like to] create the opportunity for coffee ceremony. Muslims are praying out in the open. It would be nice to accommodate this and create suitable spaces."* |
| **Toilets and Bathing Facilities** | - Insufficient resources, limited funding, and maintenance challenges contribute to sanitation and safety concerns<br>- Lack of gender-segregated toilets impacts dignity and comfort | *"The facility has no shower access. It only has toilets; we couldn't shower"* |
| **Mobility During Labour and Birth Position** | - Walking aids labour progression, but there were limited walking spaces<br>- Overcrowding and lack of grab bars create safety risks<br>- Most mothers were interested in trying alternative birth positions<br>- Providers shared physical constraints that prevented them from providing a variety of options | *"I wanted to walk around [in early labour] but there was no privacy in the [labour ward]. It was too crowded. There is so much heat in there."*<br>*"I ask if they want to deliver in another position, but I don't encourage, because I don't have the required instruments to deliver in another position."* |
| **Provider Proximity** | - Location of nurse stations can reduce or facilitate patient monitoring and emergency response<br>- Some facilities were limited to repurposing rooms that were unused for nurse stations, increasing provider-patient separation | *"I would like to see open nurses station, secured and glazed. Separate mothers room...KMC and waiting spaces."* |

### Overall Space and Adjacencies

Facility walk-throughs revealed that layouts and available space were frequently insufficient to support patient volume, privacy needs, provider workflow, and recommended care practices simultaneously. Some spaces were undersized for their intended use, while others had been repurposed in ways that limited functionality. Facilities in Bangladesh generally experienced fewer issues with unused space but faced substantial crowding and limited capacity. Facilities in Ethiopia were more likely to have unused and unsuitably repurposed spaces, in addition to limited capacity. For example, because the maternity ward at Health Centre C had originally been designed as an operating room, the postnatal room C was undersized relative to patient demand while the delivery room was disproportionately large given it accommodated only one woman at a time (see Figure 1).

**Figure 1.**
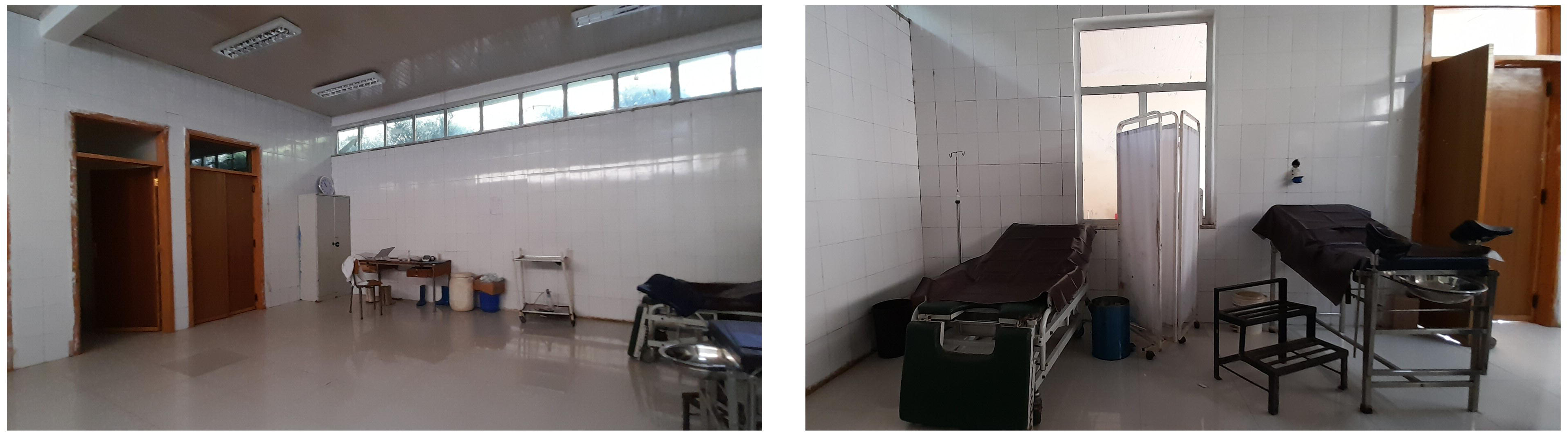
Health Centre C’s postnatal room (left) and delivery room (right) Alt text: Two side-by-side photos of Health Centre C’s postnatal room and delivery room. The postnatal room photo shows three hospital beds in a smaller space, while the delivery room photo shows one bed in a larger, emptier space.

Providers, patients, and companions all described how space limitations contributed to crowding in waiting areas, corridors, and treatment areas. Poor ventilation further intensified congestion and discomfort. Mothers across facilities expressed strong concerns about feeling vulnerable due to a lack of privacy caused by crowding, particularly when men and women were forced to share space.

> *“[There is a] lack of privacy for the delivery patient. There is no curtain outside the delivery room. That is why outsiders can see. It is very insulting for us.” (Mother, Bangladesh)*

> *“It would have been better to stop the entry of men in the ward.” (Mother, Bangladesh)*

> *“[There are] two or three patients and their attendants are present in one room. All examination and delivery is done in the same room. So, there is no privacy.” (Provider, Bangladesh)*

Inefficient layouts were also observed across multiple facilities. In Hospital D, maternal and newborn health services were dispersed across several buildings, requiring patients to navigate the confusing campus and cover long distances. In several facilities, circulation routes forced patients and visitors to pass through treatment areas to access registration desks and bathrooms, further compromising privacy and care flows.

### Companions

Mothers and providers consistently emphasized the importance of companions during labour and the postpartum period. Companions provide emotional and physical support by providing food for birthing mothers and serving as an advocate between mothers and providers, often naming the challenges mothers were unable to verbalise during labour and postpartum.

> *“Companion’s emotional support and reassurance is encouraging; they offer physical support while moving during labour.” (Mother, Ethiopia)*

> *“They gave me courage when I entered the delivery room.” (Mother, Bangladesh)*

> *“Companions can tell us problems that mothers can’t talk about.” (Provider, Bangladesh)*

However, observations through shadowing and companion interviews presented evidence that the companions’ needs were not often accommodated. Seating was limited, forcing companions to crowd treatment areas or wait in corridors and circulation spaces, contributing to congestion and privacy concerns. In Bangladesh, where women were typically accompanied by up to four people, companions frequently sat on patient beds or waited in distant areas of the facility.

> *“I have been sleeping outside the MNH block on a piece of cardboard because there is no other option while we stay here.” (Companion, Ethiopia)*

> *“There is no sitting space or even chair for companions.” (Companion, Ethiopia)*

None of the eight facilities included sleeping quarters, storage areas, cooking facilities, or laundry spaces for companions, despite many remaining at the facility for extended periods. Given the centrality of companions to the birth experience, this lack of provision for their well-being was dehumanising and was one of the factors mothers cited for choosing to leave the facility shortly after birth.

### Cultural Practices

Cultural and religious practices surrounding childbirth emerged as important components of the care experience across both countries. In Ethiopia, during labour, friends and family of the mother roast and drink coffee and burn incense during labour, companions prepare a porridge called *genfo*, which the mother eats soon after delivery, and in the Amhara Region, it is common for family members and companions to consume specific alcoholic beverages including *tella* and *areke* in celebration of the birth. In Bangladesh, cultural and religious practices surrounding childbirth include: Islamic rites performed shortly after birth, such as feeding the baby honey and performing the a*zan* (call to prayer) in the newborn’s ear as well as the Hindu practice of *ulu dhwani*. A few days after birth, Muslim families engage in a*qiqa* (sacrifices and naming ceremony), and most companions prefer to perform daily prayers.

However, facilities generally lacked designated areas for food preparation, religious observance, or cultural practices, particularly for families travelling long distances to access care. Limited access to safe and reliable water further constrained hygiene-related practices and food preparation.

> *“There should be space for cooking, reheating food, laundry facilities, and a canteen is needed for buying food.” (Mother, Bangladesh)*

Prayer and religious observance during labour and delivery were also common in both communities of Ethiopia, and stakeholders described a need for dedicated space for prayer. Hospital A and Health Centre A did include a prayer room for patients, but neither of the facilities in another district included prayer rooms.

> *“[We’d like to] create the opportunity for coffee ceremony. Muslims are praying out in the open. It would be nice to accommodate this and create suitable spaces.” (Provider, Ethiopia)*

### Toilets and Bathing Facilities

Participants identified inadequate sanitation infrastructure, maintenance and funding challenges, and limited cleaning resources as barriers to safe and dignified care. Mothers and companions described leaving facilities shortly after birth because of poor water and sanitation conditions, despite recommendations to remain for at least 24 hours postpartum.

> ***“****We couldn’t stay longer because there was no water at the facility.” (Companion, Ethiopia)*

Most facilities, like Hospital A, did not provide showers. Mothers shared how they used buckets instead to bathe. Both mothers and providers also noted how bathrooms were often wet and not regularly cleaned, raising hygiene concerns.

> *“The facility has no shower access. It only has toilets; we couldn’t shower” (Mother, Ethiopia)*

> *“The situation is not very good. The workers do as they can, but there’s only one cleaner in the ward. There is only one security guard on the first floor.” (Provider, Bangladesh)*

Participants also described inadequate numbers of functioning toilets for patients and companions. In some facilities, men and women were required to share toilets, which mothers reported as uncomfortable and undignified.

### Mobility during Labour and Alternative Birth Positions

Walk-throughs and interviews highlighted limited opportunities for mobility during across both countries. In many facilities, overcrowding and limited space prevented women from moving safely during labour or accessing designated walking areas. At Health Centre A and Health Centre C, women who wished to walk had to do so in crowded treatment areas and corridors, which created additional privacy and safety concerns.

> *“Because of the small room, mothers have to walk out of the room. That is not safe for them because there are a lot of people outside.” (Provider, Bangladesh)*

While some facilities allowed women to walk outdoors, the physical landscape made it challenging. For example, at Hospital D, the area surrounding the building was rocky, steep, polluted, and lacked sufficient lighting at night, which further discouraged mothers from moving during labour.

> *“I wanted to walk around [in early labour], but there was no privacy in the [labour ward]. It was too crowded. There is so much heat in there.” (Mother, Ethiopia)*

The absence of supportive infrastructure such as seating, grab bars, and protected walking areas further limited mobility and created safety risks for labouring women.

During interviews, most mothers in Ethiopia and Bangladesh also expressed interest in alternative birth positions. However, delivery rooms were generally not designed or equipped appropriately to support a wide range of positions. The birthing beds at Health Centre D, Hospital A, and Hospital D only supported supine positioning, while overcrowded rooms and storage of equipment within treatment spaces further constrained flexibility. Providers described how physical limitations within the delivery environment discouraged them from supporting alternative positions.

> *“We discourage mothers sometimes [from giving birth in other positions] because [the] bed design and equipment may not be appropriate for the preferred position” (Provider, Ethiopia)*

> *“I ask if they want to deliver in another position, but I don’t encourage, because I don’t have the required instruments to deliver in another position.” (Provider, Bangladesh)*

### Provider Proximity and Visibility

Facility walk-throughs and simulations identified physical barriers that limited providers’ visibility of patients in several Ethiopian facilities. For example, in Health Centre D, an antenatal room was repurposed as an ad hoc nurse station because it was one of the few spaces available, but it was further away from the patient rooms (see Figure 2). In Health Centre C, the provider spaces were located in isolated rooms with doors that were often closed, further increasing visual and auditory separation from patients. Providers in Ethiopia expressed the desire for open spaces and higher visibility within their provider spaces.

> *“I would like to see open nurses station, secured and glazed. Separate mothers room…KMC and waiting spaces.” (Provider, Ethiopia)*

**Figure 2.**
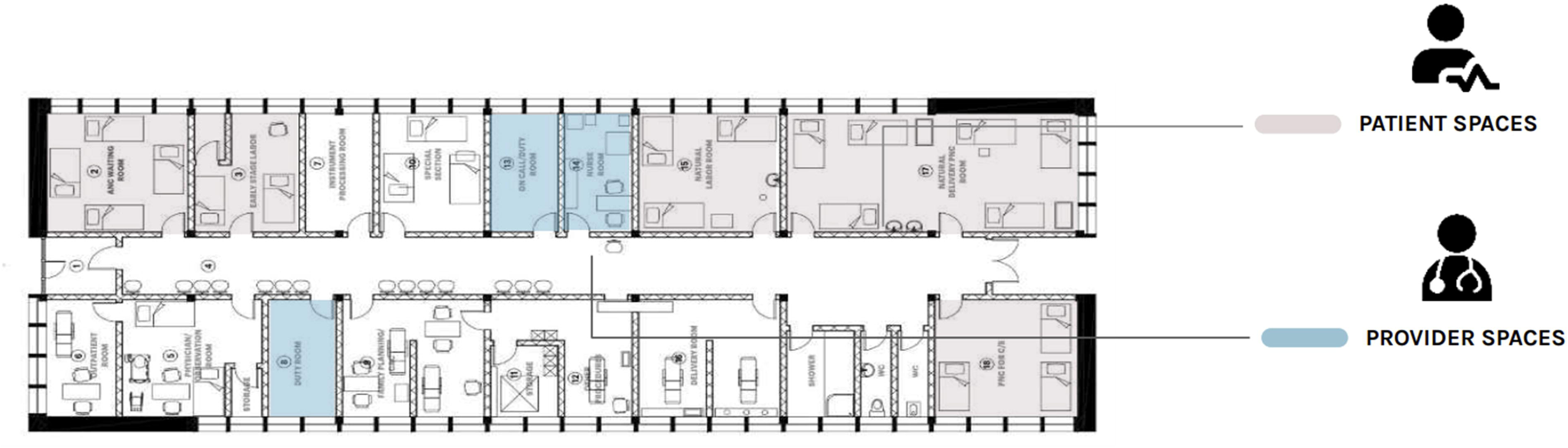
Health Centre D in Ethiopia Alt text: Floor plan diagram of Health Centre D in Ethiopia, that labels the locations of the patient and provider spaces in two different colours. The provider spaces are located farther away from patient spaces and divided by walls and private rooms.

Facilities in Bangladesh generally positioned nurse stations more centrally, improving visibility and oversight. Most wards had nurse stations adjacent to patient spaces, allowing providers to maintain audio and visual connection, while enabling nurses to complete their administrative tasks. In Hospitals A and B (see Figure 3), nurse stations were separated from the maternity wards by transparent partitions or windows, allowing providers to maintain visibility and facilitate oversight.

**Figure 3.**
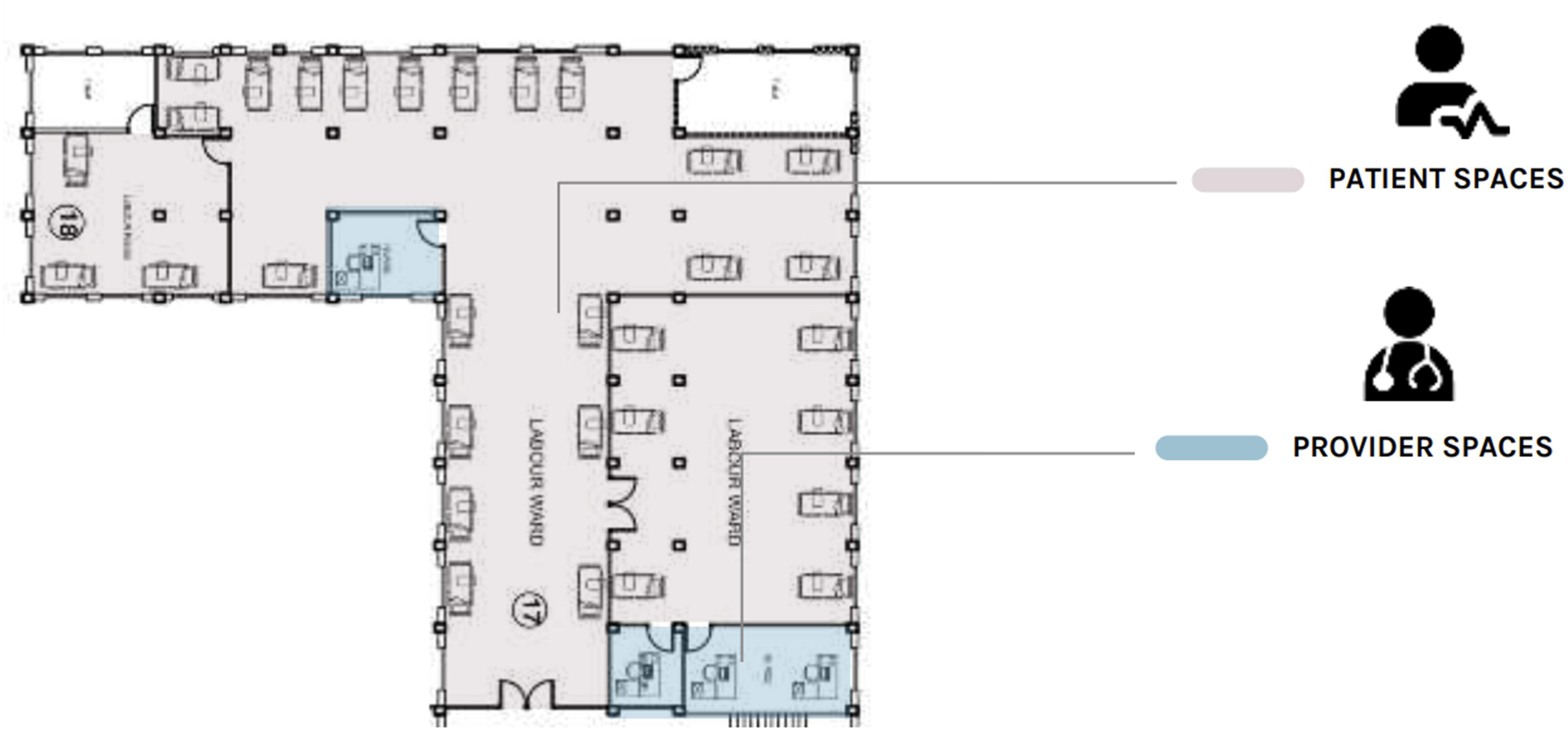
Hospital B in Bangladesh Alt text: Floor plan diagram of Hospital B in Bangladesh that labels the locations of the patient and provider spaces in two different colours. Provider spaces are located within the patient space (the labour ward), with no separation of rooms.

## Discussion

While maternal and newborn health initiatives increasingly emphasize respectful, equitable, and evidence-based care, far less attention has been paid to whether existing health facility infrastructure can adequately support these models of care. Recent studies have increasingly recognized the influence of the physical space and facility design on maternal care experiences, care-seeking behaviours, provider workflow, and health outcomes. Our study used a HCD approach to examine how the physical structure of health facilities shaped the experiences of patients, companions, and providers in Bangladesh and Ethiopia. Key focus areas included overall facility layout, companion accommodation, space for cultural and religious practices, water and sanitation facilities, mobility during labour and alternative birthing positions, and provider proximity and visibility. Based on these findings, we propose six recommendations to better align health facility infrastructure with maternal and newborn care delivery needs.

1. *Address Overcrowding:* Overcrowding is a common challenge in public health facilities across LMICs, discouraging women from seeking facility-based care during labour and delivery (Oyerinde et al., 2012). It also creates safety risks, including trip hazards and delays in providers reaching patients to deliver critical care (Hanson et al., 2020). While expanding infrastructure may not always be feasible, opportunities exist to reorganize services and improve patient flow within existing facilities. Perinatal care services should be located in adjacent spaces so that the mother can smoothly and comfortably move from one space to the next as she progresses in her birth journey. Each space should be the appropriate size for the service it offers. For example, labour and postpartum wards, which typically serve multiple mothers concurrently, should be larger than delivery rooms intended for individual births. Making greater use of outdoor space and creating dedicated waiting areas may help reduce crowding inside facilities, making mobility easier for both providers and patients actively receiving care. Reorganizing spaces and ensuring appropriate entry/exit points may also reduce intrusion into treatment areas and better protect patient privacy and comfort.
2. *Ensure Space for Companions:* In many LMICs, family and companions play a crucial role during childbirth by providing emotional support, carrying personal belongings, offering food, helping with registration, translating, and communicating with health care providers about the mother’s condition. Research shows continuous support from a birth companion is associated with improved health outcomes, shorter labour duration, more positive childbirth experiences, reduced use of pharmacological analgesia, and reduced likelihood of babies with low 5-minute Apgar scores (Bohren et al., 2017; Nilvér & Berg, 2023). Ensuring adequate space for companions may therefore improve both care experience and labour outcomes. Within treatment areas, facilities should include designated spaces for companions that allow them to support mothers without interfering with care provision, as well as space for personal belongings. Outside treatment areas, facilities should provide spaces where companions can rest, wash, and eat without disrupting patient flow or compromising privacy.
3. *Ensure Space for Cultural and Religious Practices:* Limited accommodation for cultural and religious practices may negatively impact birthing and postpartum experiences. Studies examining childbirth practices across LMICs suggest that the ability to observe traditional, spiritual, and cultural practices can influence perceptions of care and decisions about where to give birth (Aynalem et al., 2023; Aziato et al., 2016; Melesse et al., 2021). Supporting these practices within facilities may improve experiences of care and encourage women to seek care earlier in labour and remain at facilities longer following delivery. Facilities should include areas for basic food preparation and access to potable water as well as spaces for prayer or other religious or cultural practices. In some settings gender-segregated areas may also be appropriate.
4. *Address Water and Sanitation Needs:* Inadequate water and sanitation facilities within birthing facilities has been associated with maternal mortality and may contribute to avoidance of facility-based care because of perceptions of poor quality (Benova et al., 2014). Poor sanitation conditions also undermine safety, dignity, and comfort for mothers and companions during their care journey. While reliable potable water infrastructure may require bigger and longer-term investment from governments, facilities should ensure there are adequate number of functioning toilets and bathing areas for patients, companions, and providers, maintain gender-segregated sanitation facilities, and prioritize regular cleaning and maintenance.
5. *Support Labour Mobility and Alternative Birth Positions:* Mobility and upright positions during labour are associated with improved uterine activity, fetal descent, preservation of the pushing reflex, and improved uterine blood flow (De Verastegui-Martín et al., 2023). However, facilities in this study often lacked adequate space to safely support movement during labour. Dedicated, private areas adjacent to the labour wards should be provided to encourage women to walk during labour. Outdoor spaces may be appropriate, taking advantage of air circulation and sunshine, so long as they are visually and auditorily protected from other areas of the facility and include safety features such as grab bars, seating, and lighting. Facilities should also support a range of birth positions. Research has shown that the spatial-functional characteristics of birth environments can influence health outcomes, satisfaction, and empower women beyond the birthing process (Butler et al., 2020). Dynamic birth positions have been associated with reduced operative deliveries, fewer episiotomies, and shorter second-stage labour duration (Gupta et al., 2017; Lin et al., 2018; Lindgren et al., 2024; Warmink-Perdijk et al., 2016). Delivery rooms must be appropriately sized and equipped to accommodate different birth positions and related equipment, including adaptable birthing beds and birthing balls, such that mothers can practice the birth position of their choice and providers can deliver the appropriate care in those positions.
6. *Ensure Provider Proximity and Visibility:* Provider proximity and visibility are important components of patient safety, especially during emergency situations. A previous study found that locating nurse stations close to patient rooms may improve communication, oversight, and response times (Plough et al., 2019). Centrally located provider workspaces with continuous visual and auditory access to patients may improve monitoring, facilitate communication among staff, and support timely emergency response. (Plough et al., 2019). Facilities should therefore prioritize layouts that minimize unnecessary separation between providers and patients while still supporting providers’ administrative and clinical responsibilities.

### Policy implications

While clinical standards of care require the inclusion of certain spaces and equipment, too often, health facility design processes too often rely on standardized blueprints that prioritize cost and consistency with little regard for local context and community needs. Our findings suggest that these approaches may result in environments that are poorly aligned with the operational realities of delivering safe, effective and patient-centred care.

Rather than relying on standard blueprints and formulas to calculate the requisite square meterage per patient population, facilities must instead be designed according to a set of core principles (e.g., layouts that promote care flows, privacy, sanitation, mobility, natural light, and ventilation) while allowing adaptations to reflect the geographical, socio-economic, and cultural needs of the communities they serve. Our research indicates that there is an opportunity for planners in Ministries of Health and private health systems to reform the process of health facility design and renovation to incorporate meaningful engagement of both providers and patients from the outset. This approach will increase the likelihood that the resulting building reflects not only the highest standards of health care, but the everyday reality of delivering and receiving care in that space.

While design principles and user engagement will help individual facilities better meet patient and provider needs, public and private health care system leaders must also consider facility design in broader health system planning. The health care and global health industries continuously introduce new care models and interventions intended to improve maternal and newborn health outcomes; however, these initiatives rarely addressed the spatial and infrastructural conditions required to implement them effectively. Ministries of Health and implementing partners must begin to ensure that innovations in health care delivery – in both the public and private sectors – adequately address implications of health facility design and identify the minimum spatial and infrastructural requirements necessary to support evidence-based care. As health systems evaluate new interventions for potential impact, cost-effectiveness, and sustainability, they must also consider how the intervention can be introduced without contributing to crowding, privacy concerns, and barriers to patient and provider mobility.

While comprehensive in its approach, this observational study is not without limitations. Firstly, the study was limited to 4 health facilities in each country and may therefore not fully represent the diversity of conditions across Bangladesh and Ethiopia. Furthermore, the study primarily focused on health facilities in rural settings, which may differ from urban settings. Despite these limitations, our findings are in line with similar studies in other contexts and suggest that the proposed recommendations have broad relevance for improving facility-based perinatal care.

## Conclusion

This study underscores the importance of the physical environment of health facilities in shaping the experiences of mothers, companions, and providers throughout the perinatal period. By gathering insights directly from these key stakeholders, we identified several design and policy changes that may improve comfort, safety, privacy, cultural accommodations, and overall experiences of care in health facilities. Our findings suggest that both modest, low-cost adaptations and broader infrastructural investments could have a profound impact on care experience, delivery, and quality.

Integrating human-centred and participatory planning approaches into health facility design may support a shift from reactive modifications to more proactive and context-responsive infrastructure planning. This transformation is essential for creating functional spaces that serve both clinicians and their patients. Ministries of Health and health system leaders should invest in facility infrastructure and planning processes that prioritize user engagement to promote safer, more dignified, and higher quality perinatal care.

## Data Availability

All data produced in the present study are available upon reasonable request to the authors.

## Data Availability Statement

Data available on request.

